# IgG Antibodies against SARS-CoV-2 Correlate with Days from Symptom Onset, Viral Load and IL-10

**DOI:** 10.1101/2020.12.05.20244541

**Authors:** Mary K. Young, Christine Kornmeier, Rebecca M. Carpenter, Nick R. Natale, Jennifer M. Sasson, Michael D. Solga, Amy J. Mathers, Melinda D. Poulter, Xiao Qiang, William A. Petri

**Affiliations:** Department of Medicine, University of Virginia Health System, Charlottesville, VA, 22908, USA; MilliporeSigma, St. Louis, MO, 63103, USA; Department of Neuroscience, University of Virginia Health System, Charlottesville, VA, 22908, USA; UVA Flow Cytometry Core, University of Virginia, Charlottesville, VA, 22908, USA; Department of Microbiology, Immunology and Cancer Biology, University of Virginia Health System, Charlottesville, VA, 22908, USA; Department of Pathology, University of Virginia Health System, Charlottesville, VA, 22908, USA

## Abstract

The emergence of severe acute respiratory syndrome coronavirus 2 (SARS-CoV-2) has resulted in a pandemic of the respiratory disease coronavirus disease 2019 (COVID-19). Antibody testing is essential to identify persons exposed to the virus and potentially in predicting disease immunity. 183 COVID-19 patients (68 of whom required mechanical ventilation) and 41 controls were tested for plasma IgG, IgA and IgM against the SARS-CoV-2 S1, S2, receptor binding domain (RBD) and N proteins using the MILLIPLEX^®^ SARS-CoV-2 Antigen Panel. Plasma cytokines were concurrently measured using the MILLIPLEX® MAP Human Cytokine/Chemokine/Growth Factor Panel A. As expected the 183 COVID-19 positive patients had high levels of IgG, IgA and IgM anti-SARS-CoV-2 antibodies against each of the viral proteins. Sensitivity of anti-S1 IgG increased from 60% to 93% one week after symptom onset. S1-IgG and S1-IgA had specificities of 98% compared to the 41 COVID-19 negative patients. The 68 ventilated COVID-19 positive patients had higher antibody levels than the 115 COVID-19 positive patients who were not ventilated. IgG antibody levels against S1 protein had the strongest positive correlation to days from symptom onset. There were no statistically significant differences in IgG, IgA and IgM antibodies against S1 based on age. We found that patients with the highest levels of anti-SARS-CoV-2 antibodies had the lowest viral load in the nasopharynx. Finally there was a correlation of high plasma IL-10 with low anti-SARS-CoV-2 antibodies. Anti-SARS-CoV-2 antibody levels, as measured by a novel antigen panel, increased within days after symptom onset, achieving > 90% sensitivity and specificity within one week, and were highest in patients who required mechanical ventilation. Antibody levels were inversely associated with viral load but did not differ as a function of age. The correlation of high IL-10 with low antibody response suggests a potentially suppressive role of this cytokine in the humoral immune response in COVID-19.

## Introduction

Since its discovery in December 2019, SARS-CoV-2 has caused over 61.8 million cases of COVID-19 resulting in more than 1.4 million deaths (1). Disease symptoms develop between 2-14 days after virus exposure and include but are not limited to fever, cough, shortness of breath, fatigue, new loss of taste or smell, and diarrhea (2). A large proportion of infected individuals recover from the virus on their own, but some require hospitalization, supplemental oxygen and mechanical ventilation (2, 3, 4). Little is yet know about long term health effects of COVID-19 or immunity to reinfection. While polymerase chain reaction (PCR) testing for the virus is an effective way to diagnosis active infection, antibody testing is critical to identify exposed individuals and potentially predict disease timepoint and future immunity.

SARS-CoV-2 is made up of multiple proteins that the immune system can recognize as antigens. These proteins include spike protein subunits (S1 and S2), the receptor binding domain (RBD) that is found on the S1 subunit, and the nucleocapsid protein (N) enclosed in the membrane allows for determination if an individual has been exposed to the virus even if they were asymptomatic. However, there are concerns that antibodies from related coronaviruses will cross react with these tests (6, 7, 8). The relationship between time from infection and antibody production is not fully delineated nor is it understood why antibody responses have a delayed onset in some patients. As vaccines are being developed, it is important to understand what antibody responses are beneficial and promote immunity, and be able to compare antibody responses from people with natural immunity and those who have been vaccinated. The ability to quantify several antigen specific antibodies by multiplex is a valuable tool in mapping immune response. Here we describe how IgG, IgA and IgM antibody levels against SARS-CoV-2 antigens measured by the MILLIPLEX^®^ SARS-CoV-2 Antigen Panels relate with disease severity, age, days from symptom onset, viral burden and plasma IL-10.

## Methods

### Sample Collection and Study Population

Blood samples from 224 patients tested for SARS-CoV-2 by PCR between April and September 2020 were collected at the University of Virginia Medical Center. Clinical information and patient demographics were was obtained from the electronic medical records and confidentiality was maintained by assigning each patient a unique identifier. The collection of blood samples and deidentified patient information was approved by the University of Virginia Institutional Review Board (IRB-HSR #22231 and 200110). 183 of the 224 patients tested were COVID-19 positive and 41 were COVID-19 negative. Of the COVID-19 positive patients, 70 had two samples from different time points including their first available blood sample after COVID-19 testing and another 7 to 10 days later. 68 of the COVID-19 positive patients were placed on mechanical ventilation. Day of symptom onset was obtained through retrospective chart review of who tested positive for SARS-CoV-2. The start of patient’s symptoms was determined by reviewing the history of present illness from the electronic medical record. Out of 183 patients reviewed, 2 were asymptomatic for SARS-CoV-2. Of the remaining 181 patients, day of symptom onset was determined for 112 patients and was unknown for 69 patients. Nasopharyngeal SARS-CoV-2 cycle threshold (Ct) values were quantified by GeneXpert XVI and GeneXpert Infinity diagnostic systems (Cepheid, Sunnyvale, CA).

### Antibody Detection

Blood collected in EDTA was centrifuged at 1300 x g for 10 minutes, then plasma was aliquoted and stored at -80°C until testing. IgG, IgA and IgM antibody levels against SARS-CoV-2 spike protein subunits S1 and S2, RBD and N were measured in duplicate plasma samples from the 224 patients using novel MILLIPLEX^®^ SARS-CoV-2 Antigen Panel 1 IgG, SARS-CoV-2 Antigen Panel 1 IgA and SARS-CoV-2 Antigen Panel 1 IgM (Millipore Sigma, St. Louis, MO, Catalog Numbers: HC19SERG1-85K, HC19SERA1-85K, and HC19SERM1-85K respectively; For Research Use Only. Not For Use In Diagnostic Procedures). This panel is designed to measure antibodies by median fluorescent intensity (MFI). The four antigens are recombinant poly-his-tagged. Samples were diluted 1:100 in assay buffer. 96-well plates were pre-wetted with 200 µL wash buffer, covered with plate sealer and incubated for 10 minutes at room temperature with shaking, then emptied. 25 µL of each diluted sample was added to the sample wells and 25 µL of assay buffer was added to background wells. 60 µL of both sonicated (30 seconds) and vortexed (1 minute) analyte and control bead was combined and brought to a final volume of 3 mL with the addition of assay buffer, vortexed, and 25 µL of bead mixture was dispensed into each plate well. The plate was sealed and incubated for 2 hours at RT with constant shaking. A handheld magnetic plate washer was used to retain magnetic beads while liquid contents were discarded appropriately, and wells were washed 3 times with 200 µL wash buffer. 50 µL of phycoerythrin-anti-human immunoglobulin (IgG, IgA or IgM per kit in use) detection antibody was added to each well, plate sealed and incubated 90 minutes at RT with constant shaking. Plates were washed three more times with magnetic plate washer. 150 µL Sheath Fluid was added to each well, the plate was then sealed and shaken at RT for 5 minutes. The plate was then read on a Luminex^®^ MAGPIX™Instrument System with a minimum of 50 beads of each analyte collected per well.

### Il-10 Detection

Il-10 in plasma were measured using the MILLIPLEX® MAP Human Cytokine/Chemokine/Growth Factor Panel A (48 Plex) (Millipore Sigma, St. Louis, MO, Catalog Number HCYTA-60K-PX48, For Research Use Only. Not For Use In Diagnostic Procedures).

### Statistical Methods

All statistical comparisons and graphs were made using GraphPad Prism 8 software. Mann-Whitney U tests were performed to compare initial antibody levels between COVID-19 positive and negative groups and different age groups of COVID-19 positive patients. Sensitivity and specificity were calculated in GraphPad Prism. Simple linear regression and Spearman correlations were used to associate antibody levels with days from symptom onset in COVID-19 and assess the relationship between viral load and IgG antibodies that are specific for SARS-CoV-2 antigens. Patient’s with CT values of zero were excluded from analysis. A non-linear regression analysis with the y= log(x) function was performed in R Studio to correlate IL-10 levels from initial samples with IgG levels in ventilated and not ventilated COVID-19 positive patients (Not Ventilated n=40; Ventilated n = 51). A p value <0.05 was considered statistically significant.

## Results

### Antibody Response to SARS-CoV-2 in COVID-19 Positive and Negative Patients

A total of 224 patients were tested for IgG, IgA and IgM antibodies against SARS-CoV-2 S1, S2, RBD and N proteins. Of these patients, 183 were positive for COVID-19 and 41 were negative. 68 of the COVID-19 positive patients were ventilated and 115 were not. COVID-19 positive patients had significantly higher antibodies against all SARS-CoV-2 proteins compared to COVID-19 negative patients (Figure 1, Supplemental Figure 1). Specificity was high for all antigens, specifically S1-IgG and S1-IgA had specificities of 97.6% and S1-IgM that had a specificity of 92%. IgA antibodies against all antigens were elevated in COVID-19 positive ventilated patients compared to not ventilated COVID-19 positive patients. IgG antibodies against S1, S2 and RBD were significantly increased in ventilated patients compared to not ventilated COVID-19 positive patients, and antibodies against N were trending higher in ventilated patients. IgM antibodies against S1, S2 and N were also significantly higher in ventilated individuals (Figure 1, Supplemental Figure 1).

**Figure 1:**
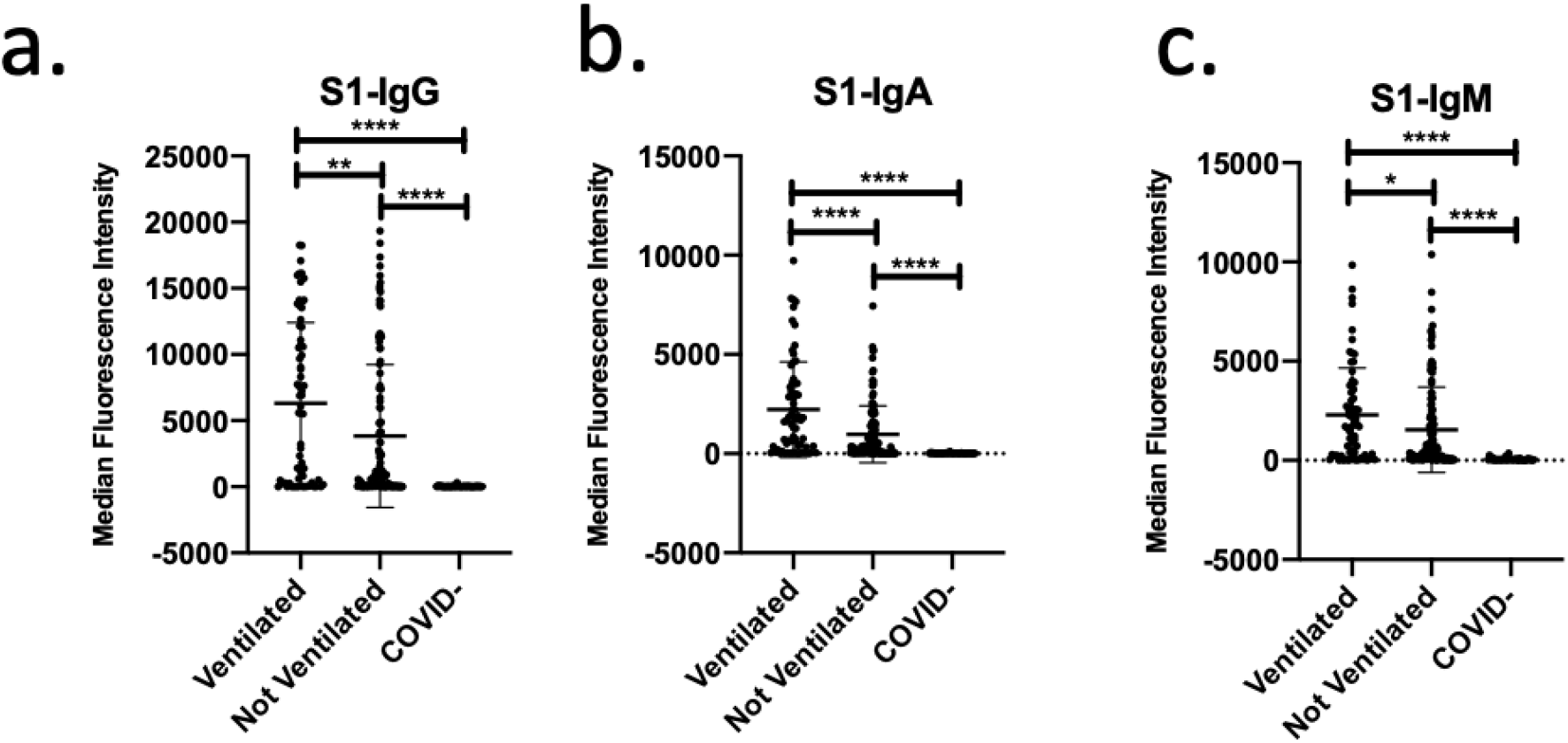
IgG, IgA and IgM antibody response to SARS-CoV-2 S1 increased in ventilated patients. (a-c) IgG, IgA and IgM antibody responses to SARS-CoV-2 S1 in ventilated COVID-19 positive patients (n=68), not ventilated COVID-19 positive patients (n=115), and COVID-19 negative patients (n=41). ****p<0.0001, ***p<0.001, **p<0.01, *p<0.05

### Antibody Response to SARS-CoV-2 in COVID-19 Positive Patients and Age

COVID-19 positive patients were divided into 4 age groups (<30, 30-49, 50-69 and >70 years old) and their antibody levels were compared. There were no statistically significant difference in IgG, IgA and IgM antibodies against S1 between the different age groups (Figure 2, Supplemental Figure 2).

**Figure 2:**
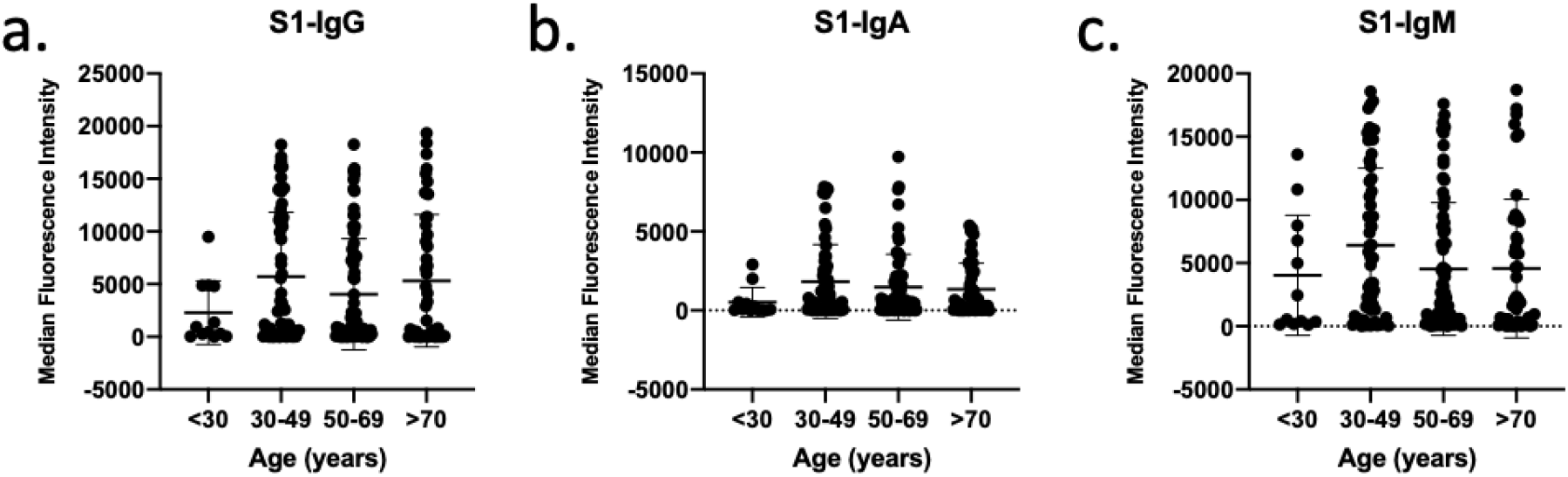
IgG, IgA and IgM antibody response to SARS-CoV-2 S1 and age. (a-c) IgG, IgA and IgM antibody responses to SARS-CoV-2 S1 in patients less than 30 years old (n=12), 30-49 years old (n=53), 50-69 years old (n=70) and greater than 70 years old (n=47).

### Correlation of Antibody Levels and Days from Symptom Onset

Antigen-specific antibodies were analyzed as a function of days from symptom onset (Supplemental Figure 3a-c). All correlations were statistically significant. IgG antibodies against S1 were most positively correlated with days from symptom onset with an r^2^ value of 0.4030 compared to IgA (r^2^=0.2142) and IgM (r^2^=0.2658) antibodies (Figure 3a-c). IgG antibodies against RBD and S2 followed a similar pattern of correlation as antibodies S1 (Supplemental Figure 3a). Sensitivity also went up after one week from symptom onset. S1-IgG went from 59.6% sensitivity to 92.5%, S1-IgA from 66% to 93.3% and S1-IgM from 68.1% to 95.8%.

**Figure 3:**
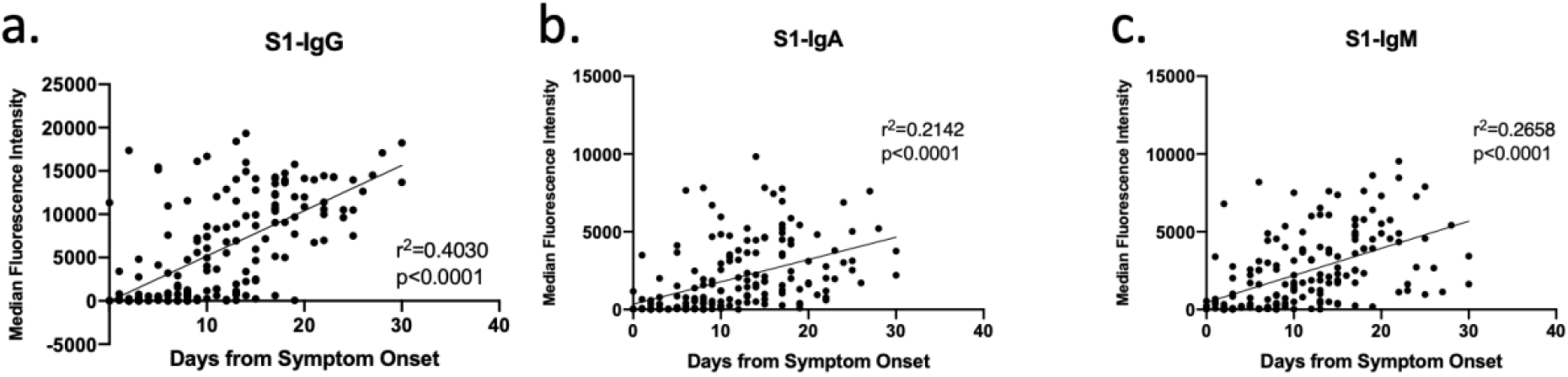
Correlation of IgG, IgA and IgM antibodies against SARS-CoV-2 S1 and days from symptom onset. (a-c) Correlation of IgG, IgA and IgM antibodies against SARS-CoV-2 S1 and days from symptom onset (168 samples from 123 patients).

### Correlation of IgG Antibody Levels and Viral Load

IgG antibody levels were correlated to clinical Ct values. IgG antibodies against S1, S2, RBD and N were found to be positively correlated with Ct values, indicating that patients with lower viral titers have higher levels of IgG (Figure 4).

**Figure 4:**
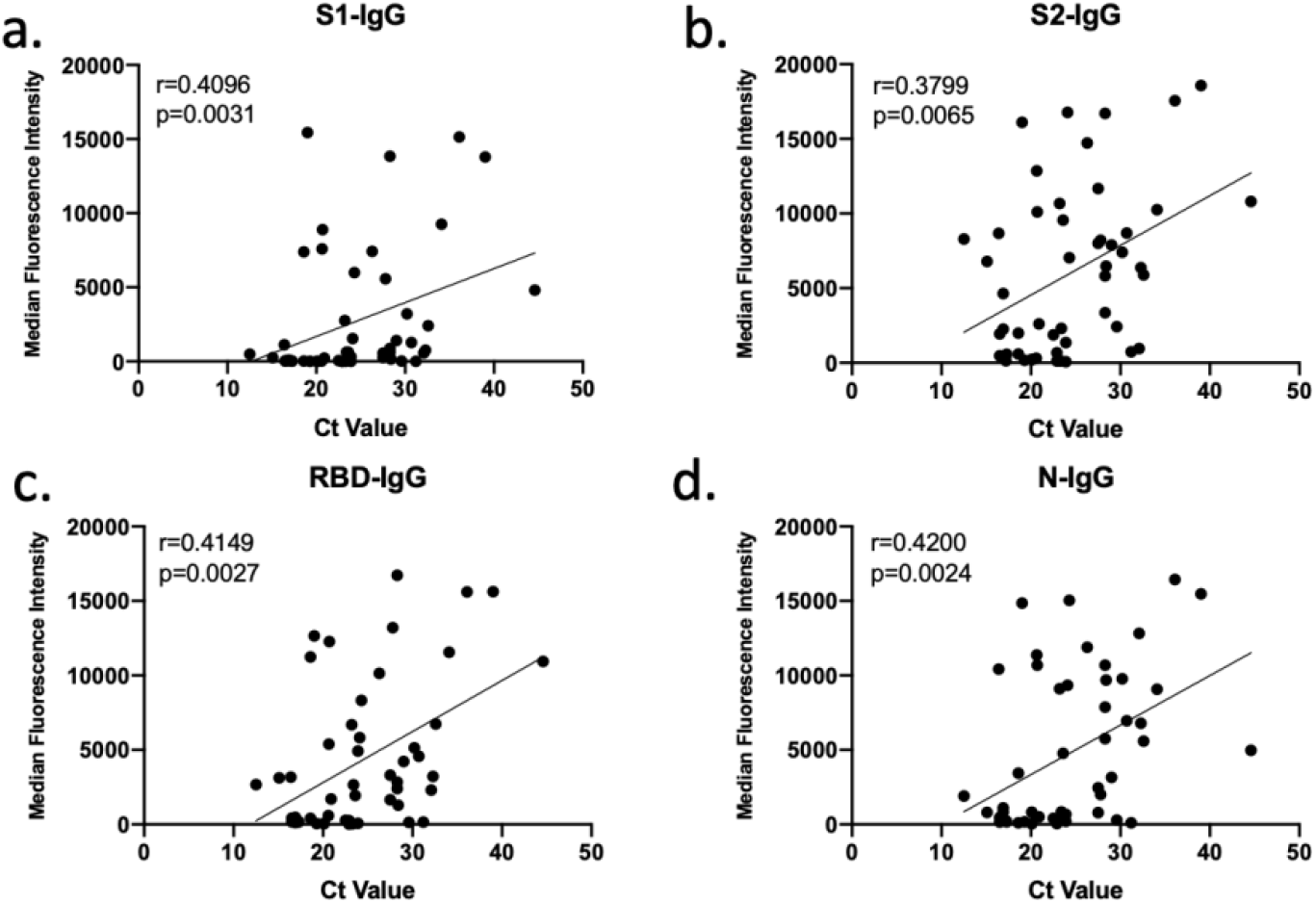
Correlation of IgG Antibodies and SARS-CoV-2 Ct Value. (a-d) Correlation of IgG antibodies against SARS-CoV-2 S1, S2, RBD and N and SARS-CoV-2 Ct Value (n=50).

### Correlation of IgG Antibody Levels and Il-10

IgG antibody levels were correlated to Il-10 levels. Anti-S1, S2, RBD and N IgG antibodies were found to negatively correlate to Il-10 in COVID-19 positive patients who received mechanical ventilation (Figure 5).

**Figure 5:**
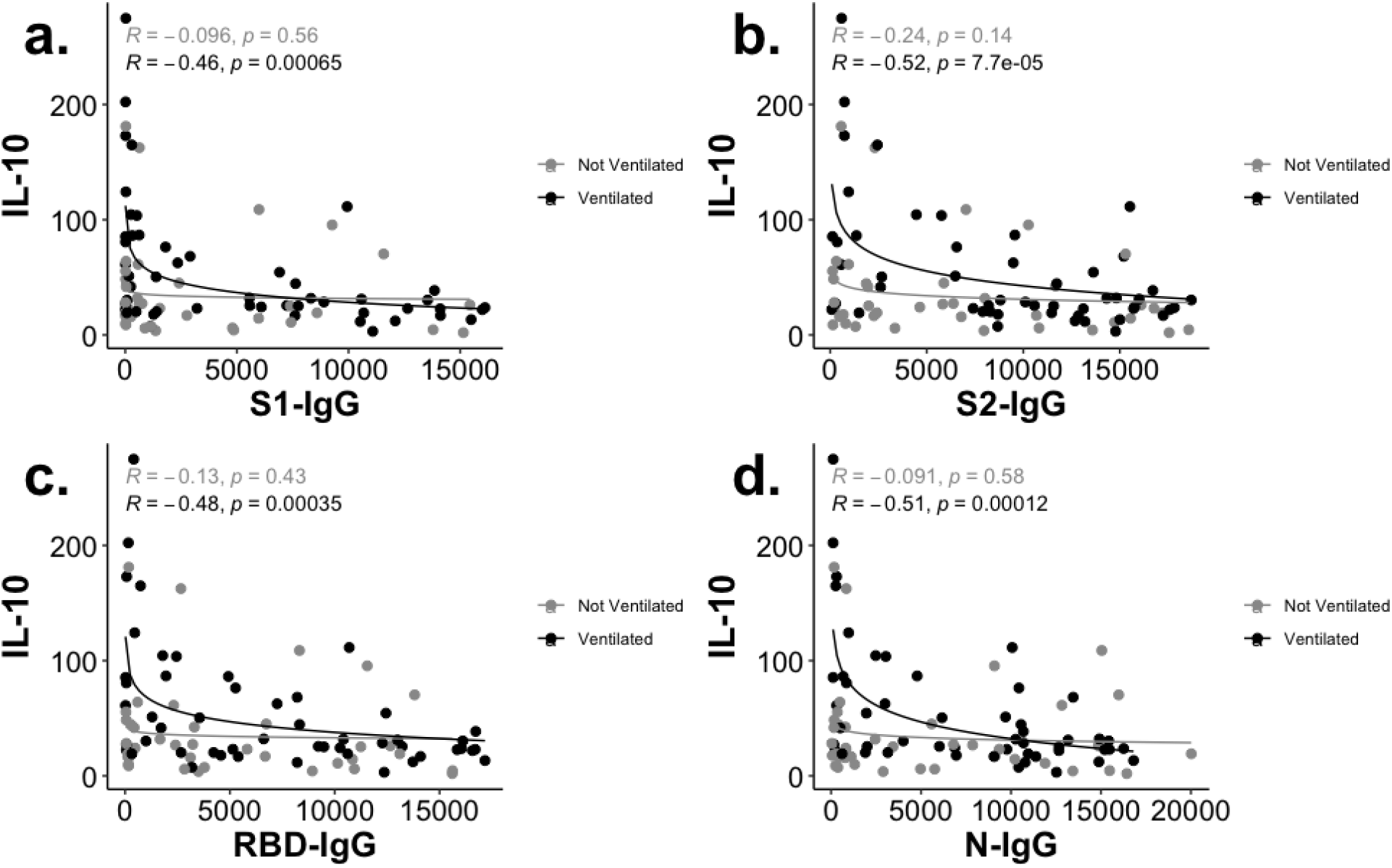
Correlation of IgG Antibodies and Il-10. (a-d) Correlation of anti-S1, S2, RBD and N IgG antibodies and Il-10 in ventilated (black, n=51) and not ventilated (grey, n=40) COVID-19 positive patients.

## Discussion

The development of accurate serological testing is critical during the COVID-19 pandemic to efficiently determine exposure to SARS-CoV-2. Here we demonstrated sensitive and specific detection of IgG, IgA and IgM antibodies against SARS-CoV-2 antigens S1, S2, RBD in COVID-19 positive patients. There was little apparent cross-reactivity with other related coronaviruses with the exception of IgG against S2 which showed modest reactivity in COVID-19 (-) patients, alleviating concerns of false positive antibody tests (7, 8). Additionally, ventilated COVID-19 positive patients had statistically significant higher antibody levels against most antigens compared to not ventilated COVID-19 positive patients. This confirms similar findings that individuals with more severe disease have higher antibody levels (9, 10, 11, 12, 13, 14). Further studies need to be done to understand the relationship between increased antibody production and ventilation.

Age has been shown to be the biggest risk factor for more severe disease and death due to COVID-19. Being over 50 doubles the risk of mortality and over 80 has a 20-fold increase risk of death (15). Here we have shown there are no significant differences in antibody levels, suggesting that antibody production does not contribute to age-related mortalities. We were able to determine days from symptom onset for 112 of the 181 COVID-19 positive patients and of those 45 patients had longitudinal samples 7 to 10 days after their initial samples. We correlated antibody levels in all of these patient samples with days from symptom onset. IgG antibodies best correlated with time from symptom onset. IgA and IgM antibodies did significantly increase over time, but had a weaker coorelations compared to IgG. This suggests that measuring IgG levels can help predict where a patient may be in their disease course. Sensitivity also went up with time from symptom onset, with all antibodies nearing 100% sensitivity after one week. Other researchers have detected antibodies present as early as 2-4 days after symptom onset with all patients producing antibodies by 14 days, similar to what we found (14, 16). Ng et al. found that individuals not infected with SARS-CoV-2, particularly children and young adults, have anti-S2 antibodies that are linked to other human coronavirus (17). In Supplemental Figure 3 A, we demonstrate that S2-IgG antibodies are present in patients as early as the day of symptom onset, suggesting that these antibodies may be boosted from a previous coronavirus infection. While anti-S2 antibody levels are significantly higher in patients with COVID-19, there are several patients with no prior SARS-CoV-2 infection that have these antibodies (Supplemental Figure 1).

Studies have indicated that higher SARS-CoV-2 viral burden results in increased disease severity (18, 19). Here we correlated antibody levels with threshold cycle values (Ct values) from initial COVID-19 diagnosis and found that IgG antibodies positively correlated with Ct values. This suggests that patients have higher antibody responses have lower viral burden. Wang et al found similar results when comparing Ct values to antibody titers (20). These results could suggest that patients with stronger antibody response are able to clear the infection better. This may also be indicative of patients being tested further from symptom onset and therefore having lower viral burden and higher antibody levels. IgG antibodies also negatively correlated with Il-10 levels. Activation of the Il-10 receptor on B cells has been reported to promote B cell survival and differentiation into IgM and IgG secreting plasmablasts. The association of high IL-10 with low antibody responses in ventilated patients is therefore apparently paradoxical, and worthy of further study (21).

To conclude, we found that the MILLIPLEX^®^ SARS-CoV-2 Antigen Panels successfully detected antigen specific antibodies in patients with COVID-19 and that patients who needed mechanical ventilation had higher IgG, IgA and IgM antibodies compared to not ventilated patients. While some antibody levels are lower in patients under 30, we did not see a strong correlation between age and antibody levels. We did find that IgG better correlates with days from symptom onset compared to IgA and IgM antibodies. With the approaching availability of COVID-19 vaccinations, this test would also be beneficial in determining whether a person has immunity due to natural infection or immunity from vaccination. Vaccinated individuals would potentially have titers against spike proteins but not the nucleocapsid. These results indicate the importance of antibody testing to determine disease time point and potential predict disease severity. This multiplex assay will also be beneficial in mapping immune response to predict potential immunity.

## Data Availability

The data will be available online when the peer-reviewed manuscript is published.

## Acknowledgements

We would like to thank Pan Tongvichit, the University of Virginia Biorepository and Tissue Research Facility, and iTHRIV for their help collecting patient samples and clinical information. This research was funded by the Manning Family Foundation, the Ivy Foundation COVID-19 Translational Research Fund and NIH grant R01 AI124214.

