## Supplemental Figures for "IgG Antibodies against SARS-CoV-2 Correlate with Days from Symptom Onset, Viral Load and IL-10"

**Supplemental Figure 1.**

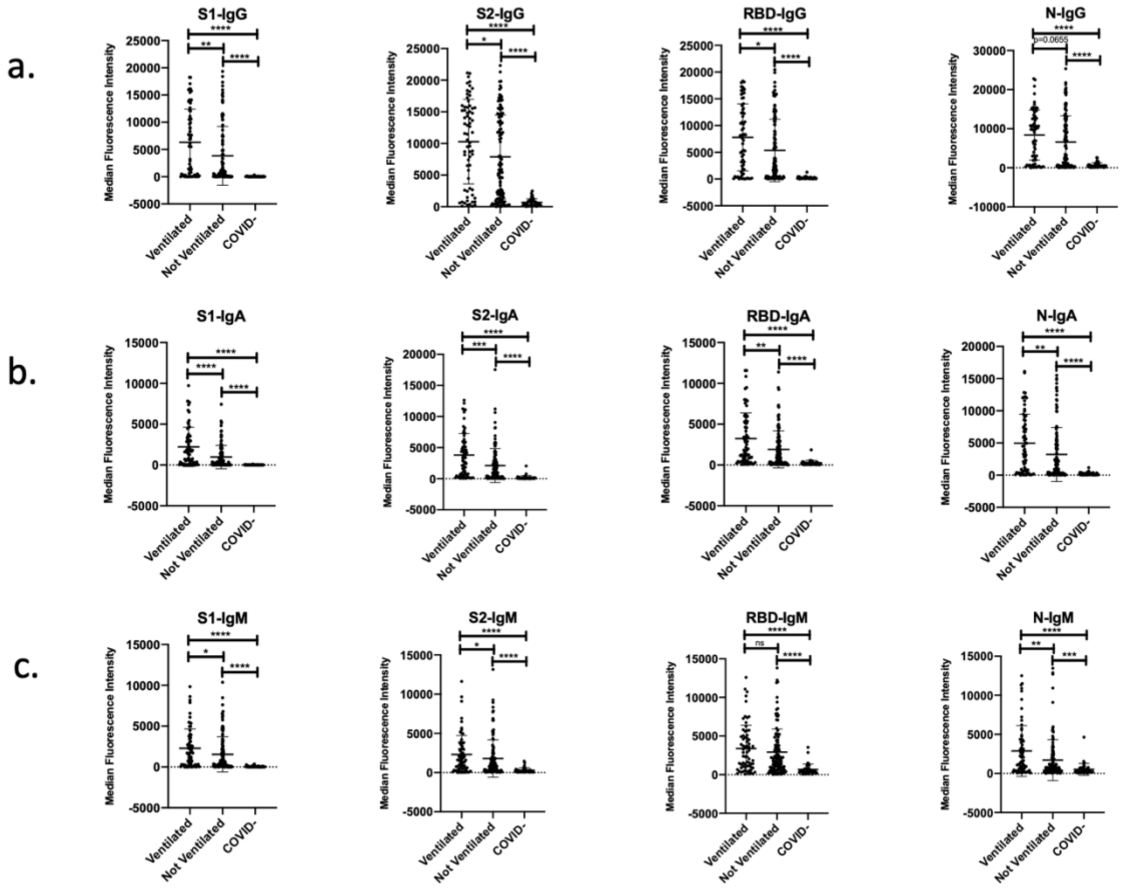

**Supplemental Figure 2.**

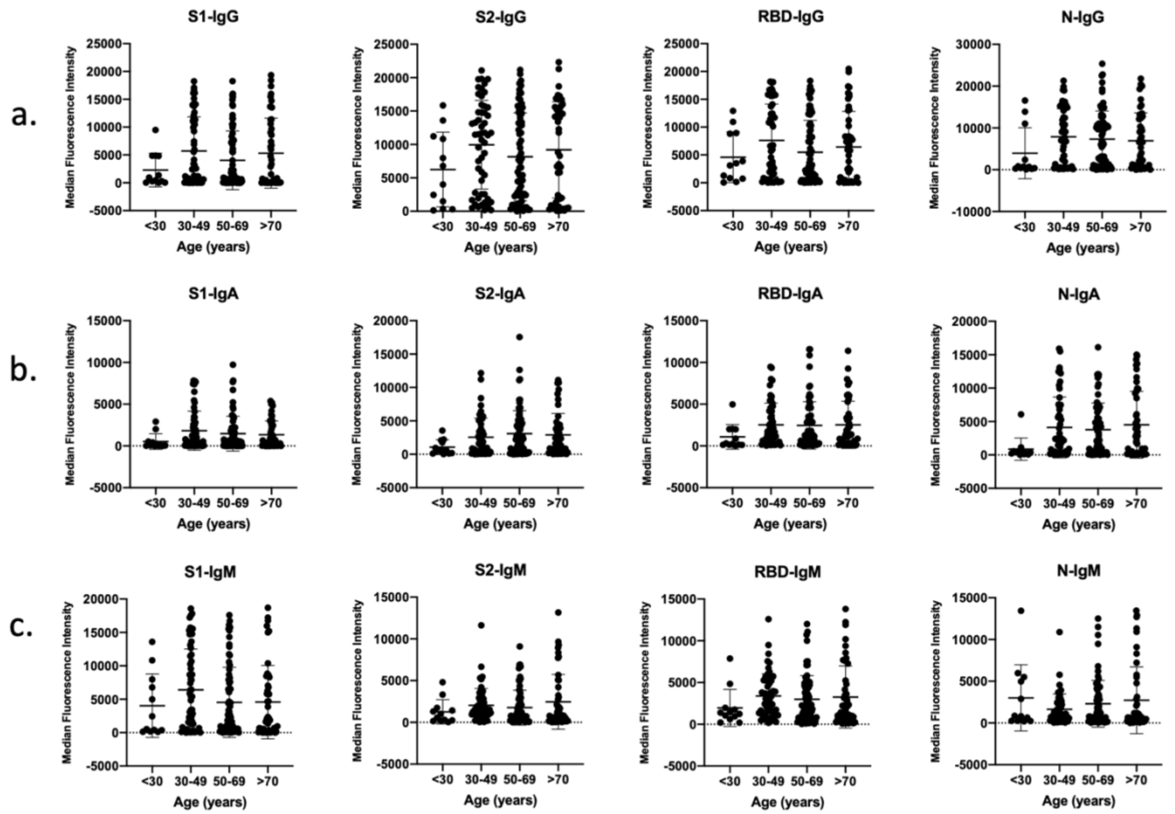


**Supplemental Figure 3.**

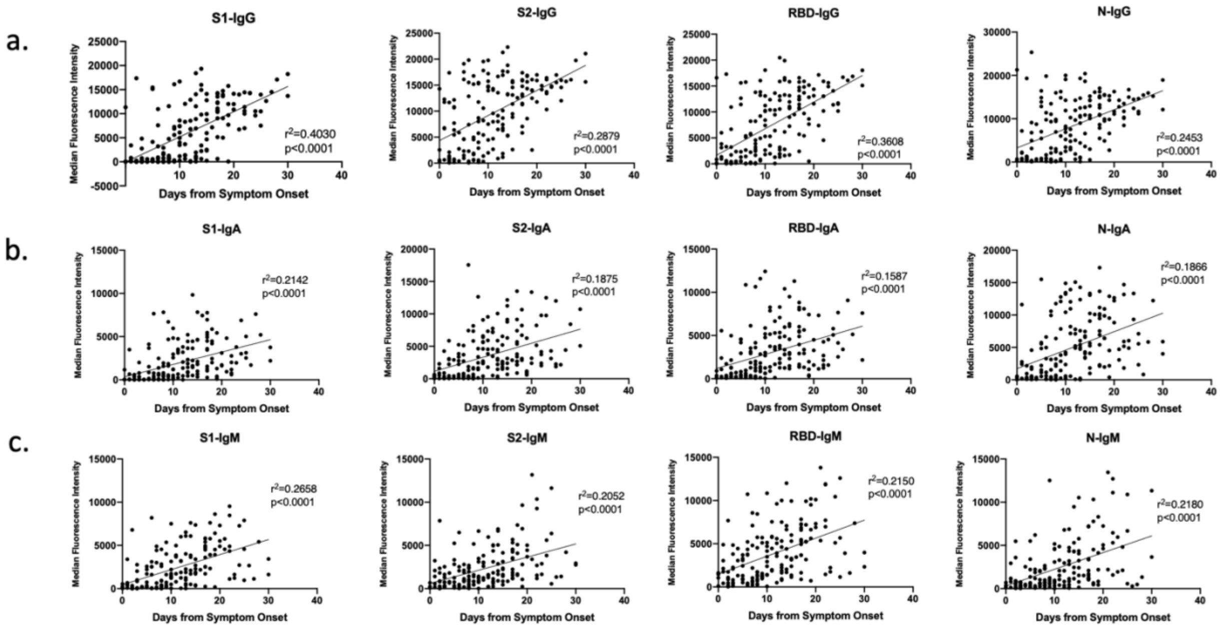

**Supplemental Figure 1: IgG, IgA and IgM antibody response to SARS-CoV-2 antigens increased in ventilated patients.** (a-c) IgG, IgA and IgM antibody responses to SARS-CoV-2 S1, S2, RBD and N in ventilated COVID-19 positive patients (n=68), not ventilated COVID-19 positive patients (n=115), and COVID-19 negative patients (n=41). ****p<0.0001, ***p<0.001, **p<0.01, *p<0.05

**Supplemental Figure 2: IgG, IgA and IgM antibody response to SARS-CoV-2 and age.** (a-c) IgG, IgA and IgM antibody responses to SARS-CoV-2 S1, S2, RBD and N in patients less than 30 years old (n=12), 30-49 years old (n=53), 50-69 years old (n=70) and greater than 70 years old (n=47).

**Supplemental Figure 3: Correlation of IgG, IgA and IgM antibodies against SARS-CoV-2 antigens and days from symptom onset.** (a-c) Correlation of IgG, IgA and IgM antibodies against SARS-CoV-2 S1, S2, RBD and N and days from symptom onset (168 samples from 123 patients).
